# DNA methylation profiling of immune cells from tuberculosis-exposed individuals overlaps with BCG-induced epigenetic changes and correlates with the emergence of anti-mycobacterial ‘corralling cells’

**DOI:** 10.1101/2021.09.01.21262945

**Authors:** Isabelle Pehrson, Clara Braian, Lovisa Karlsson, Nina Idh, Eva Kristin Danielsson, Blanka Andersson, Jakob Paues, Jyotirmoy Das, Maria Lerm

## Abstract

The mechanism of protection of the only approved tuberculosis (TB) vaccine, Bacillus Calmette Guérin (BCG) is poorly understood. In recent years, epigenetic modifications induced by BCG have been demonstrated to reflect a state of trained immunity. The concept of trained immunity is now explored as a potential prevention strategy for a variety of infections. Studies on human TB immunity are dominated by those using peripheral blood as surrogate markers for immunity. Here, we instead studied the lung compartment by obtaining induced sputum from subjects included in a TB contact tracing. CD3- and HLA-DR-positive cells were isolated from the collected sputum and DNA methylome analyses performed. Unsupervised cluster analysis revealed that DNA methylomes of cells from TB-exposed individuals and controls appeared as separate clusters, and the numerous genes that were differentially methylated were functionally connected. The enriched pathways were strongly correlated to previously reported epigenetic changes and trained immunity in immune cells exposed to the BCG vaccine in human and animal studies. We further demonstrated that similar pathways were epigenetically modified in human macrophages trained with BCG *in vitro*. Finally, we found evidence of an *M. tuberculosis-*triggered emergence of a non-macrophage cell population from BCG-trained macrophage cultures. These cells did not phagocytose *M. tuberculosis*, but ‘corralled’ the bacteria into focal points, resulting in limitation of bacterial growth. Altogether, our study demonstrates that similar epigenetic changes are induced by *M. tuberculosis* and BCG and suggests that the modifications promote transformation of macrophages (or an unknown progenitor) to establish a yet undescribed cellular defense mechanism which we term ‘corralling’, based on the metaphorical resemblance to sheepdog herding.

## Introduction

Tuberculosis (TB) is a pulmonary infection of pandemic rank and an expansion of the current toolkit for diagnosis, prevention and treatment is critical for reaching the United Nations’ Sustainable Development Goals for 2030 of ending the TB epidemic^1^. TB is caused by *Mycobacterium tuberculosis*, which transmit via aerosols and target alveolar macrophages in exposed individuals^2^. In susceptible hosts, the bacteria replicate inside the macrophages and use them as trojan horses in order to disseminate in the tissues ^3^. Bacillus Calmette Guérin, a non-virulent derivative of *Mycobacterium bovis*, has been used for almost a century as a vaccine against TB, with variable efficacy. Numerous studies have failed to provide correlates of protection, leaving the vaccine mechanism elusive. In recent years, the concept of trained immunity has evolved as an epigenetically encoded immune memory that can be triggered by a variety of stimuli and is reflected in a reprogrammed immune state characterized by a higher magnitude of response to subsequent pathogen challenges. The discovery of epigenetically regulated antimicrobial defense mechanisms goes beyond the classical understanding of immune defense and opens up a new field of research. Along this line, we have demonstrated that administration of the BCG vaccine to healthy subjects induced profound epigenetic alterations in immune cells, which correlated with enhanced anti-mycobacterial activity in macrophages isolated from the vaccinees^4^. The changes were reflected in the DNA methylome, with the strongest response being recorded within weeks after vaccination^4^. Our observation that BCG induces alterations of the DNA methylome of immune cells has later been confirmed by others^5,6^. Since BCG vaccination reflects an *in vivo* interaction between immune cells and viable mycobacteria, we here hypothesized that natural exposure to *M. tuberculosis* would induce similar changes not only in TB patients, but also in individuals who have been exposed to TB. Analyses of DNA methylomes of immune cells isolated from lungs and peripheral blood allowed us to identify distinct DNA methylation (DNAm) signatures in TB-exposed individuals. Pathway analyses revealed strong overlaps with previous studies on BCG-induced epigenetic signatures that could be correlated with protection against *M. tuberculosis*. We therefore set up an experiment to scrutinize the anti-mycobacterial mechanisms correlating with BCG-induced epigenetic reprogramming in human primary macrophages. BCG-trained macrophages were subjected to DNA methylome analysis and in parallel monitored in a live cell imaging system during exposure to *M. tuberculosis*. Again, the DNAm change *in vitro* highly overlapped with the signatures of the TB-exposed individuals and the previously identified BCG-induced epigenetic changes. Guided by these findings, we designed experiments to monitor the events taking place in BCG-trained cells challenged with *M. tuberculosis*. To our surprise, rapidly dividing, non-phagocytic cells with a size of approximately 1/4^th^ of the diameter of macrophages appeared in BCG-trained, *M. tuberculosis-*infected macrophage cultures. These ‘corralling cells’, whose nature remains elusive, apparently concentrated the bacteria into focal points by pushing them in a coordinated fashion, while limiting their growth. To our knowledge, this phenomenon has not been described previously.

## Results

### Study design

To determine epigenetic changes in the immune cells in TB-exposed individuals, we recruited subjects enrolled in a routinely performed TB contact tracing at Linköping University Hospital, Sweden. Age-matched individuals were included as controls (Table 1). The index case was diagnosed with drug-sensitive pulmonary TB and had completed two out of six months of standard treatment at the time of sample collection. All included subjects except one (a TB contact) had been BCG-vaccinated previous to the study (Table 1). Interferon-Gamma Release Assay (IGRA) status was determined and among the exposed individuals, two were positive (including the index case) and among the controls, one individual (C2) was classified as ‘borderline’- positive^7^ (Table 1). From induced sputum, we used an established protocol for the isolation of HLA-DR-positive antigen-presenting cells, dominated by macrophages^8^ and T cells (CD3 positive cells) (BioRXiv, 2021, pre-print, accepted for publication in *Epigenetics*), whereas the PBMC fraction extracted from blood was kept as a mixed population. For the in vitro experiment with BCG and *M. tuberculosis* exposure, human primary monocytes were collected from blood samples from healthy blood donors and differentiated into macrophages. (Fig. 1).

**Table 1:** Demographic data of the participants. ^†^The standard deviation of the mean values is added to the age, height, weight and BMI. ^*^ Borderline-positive.

| Characteristics | TB-exposed (n = 4) | Controls (n = 6) |
| --- | --- | --- |
| Mean age (year)<sup>†</sup> | 36±12 | 27±6 |
| Mean height (cm)<sup>†</sup> | 173±4 | 176±7 |
| Mean weight (kg)<sup>†</sup> | 69±8 | 77±16 |
| Mean Body Mass Index (BMI)<sup>†</sup> | 23±3 | 24±5 |
| Sex (male/female) | 1/3 | 4/2 |
| Smoking (current/previous/never) | 0/1/3 | 0/0/6 |
| BCG (yes/no) | 3/1 | 6/0 |
| IGRA-positive/IGRA-negative | 2/2 | 1*/5 |

**Figure 1.**
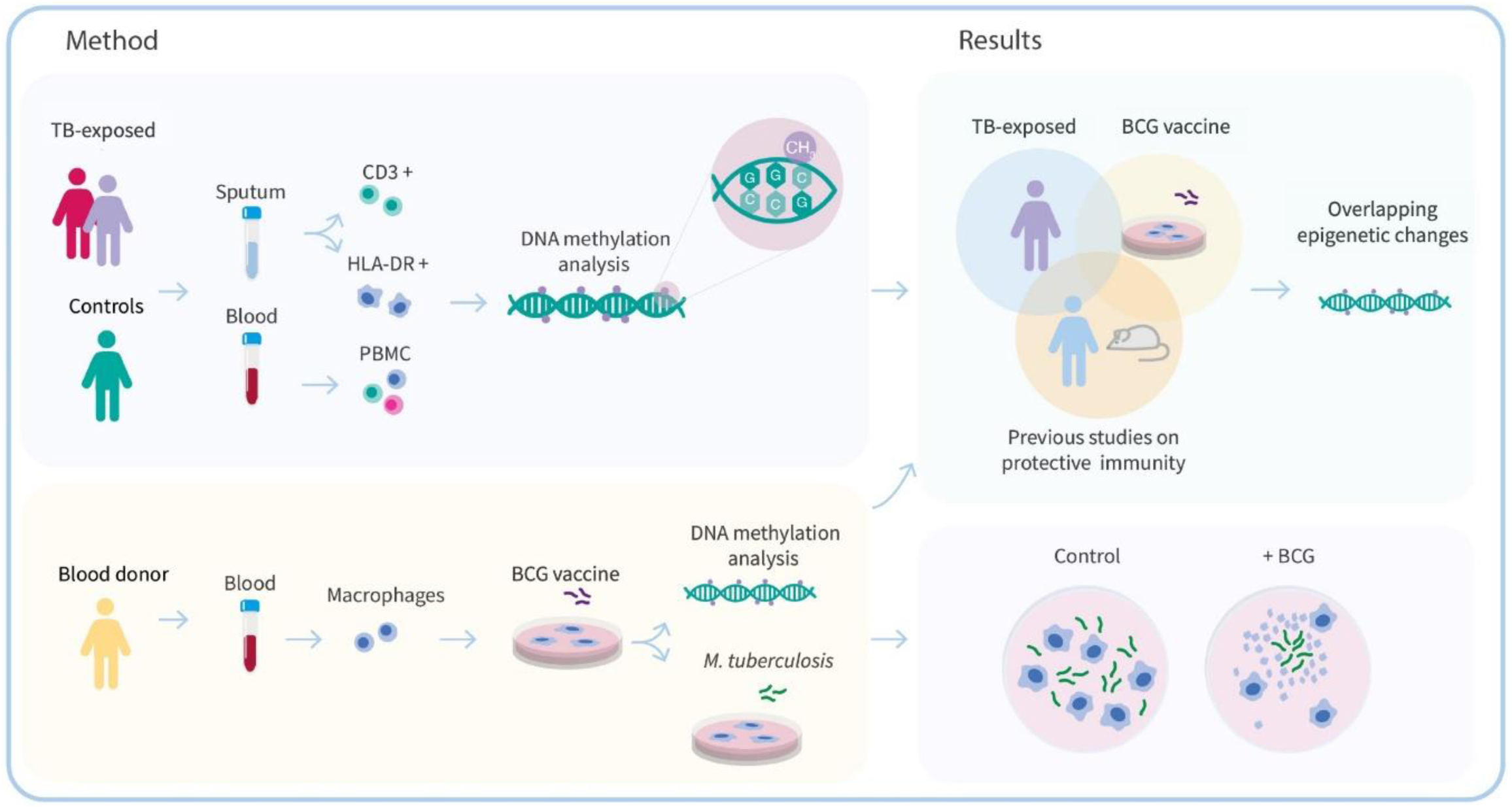
Schematic overview of the project workflow. Sputum and blood were isolated from TB-exposed individuals and controls and the DNA methylomes were analyzed for the different cell types. Macrophages were isolated from healthy blood donors and exposed to the Bacillus Calmette–Guérin (BCG) vaccine *in vitro*. The BCG-exposed macrophages were subjected to DNA methylome analysis and in parallel, infected with *M. tuberculosis* and monitored in a live cell imaging system.

### DNA methylome data from TB-exposed individuals form a separate cluster

DNA isolation from the studied cell populations was followed by global DNAm analysis using the Illumina 450K protocol. After curation of the data^9^, the datasets were subjected to unsupervised hierarchical cluster analysis based on DNA CpG methylation β-values. This approach accurately clustered the participants into TB-exposed and controls based on the DNA methylome data derived from both HLA-DR- and CD3-positive cell populations. (Fig. 2a, b). On the other hand, in the PBMC-derived dataset, the TB index case appeared outside the clusters and two of the controls clustered with the other exposed individuals, one of them (“Con_2”) being the individual identified as borderline-positive in the IGRA test (Fig. 2c).

**Figure 2.**
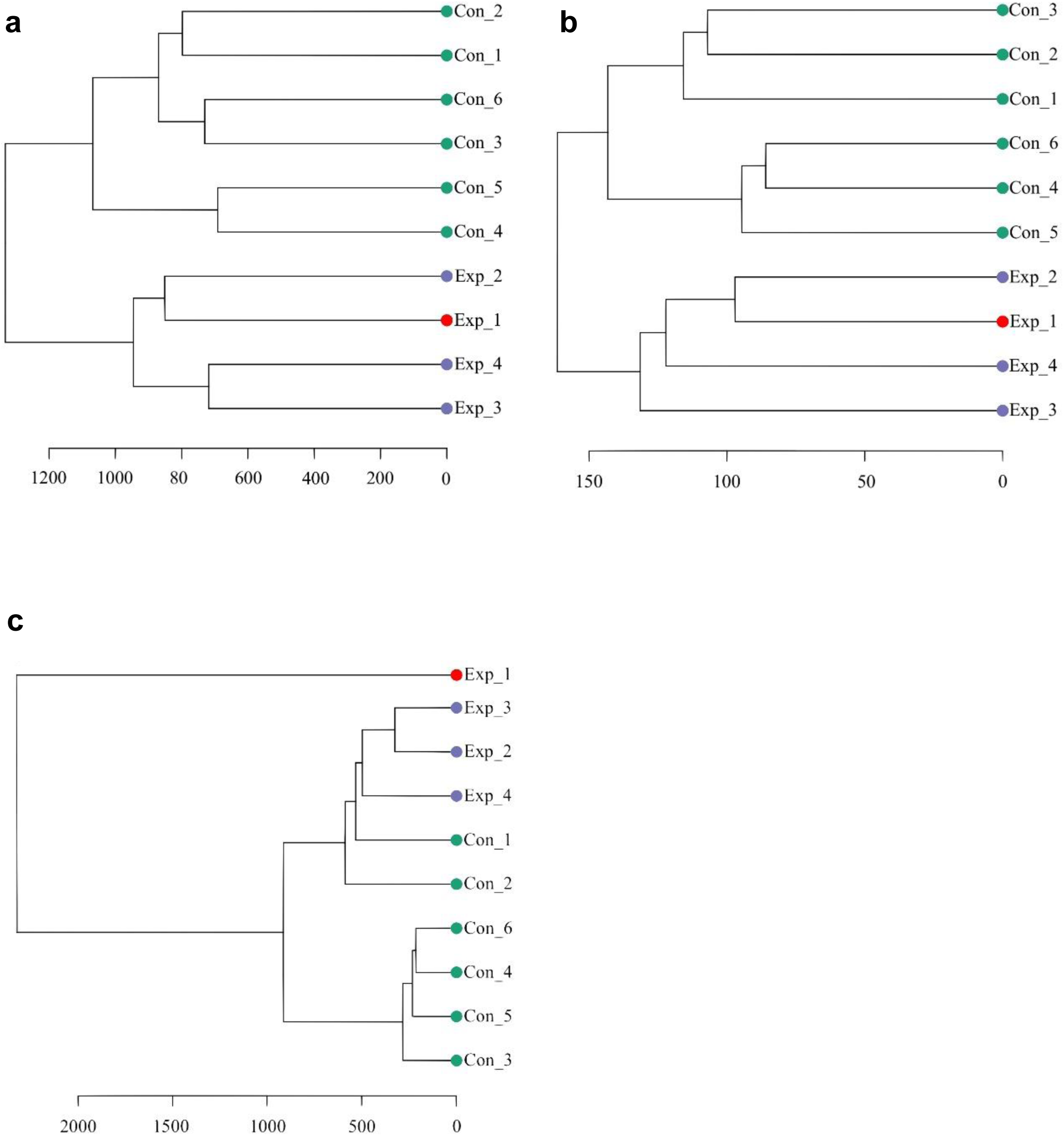
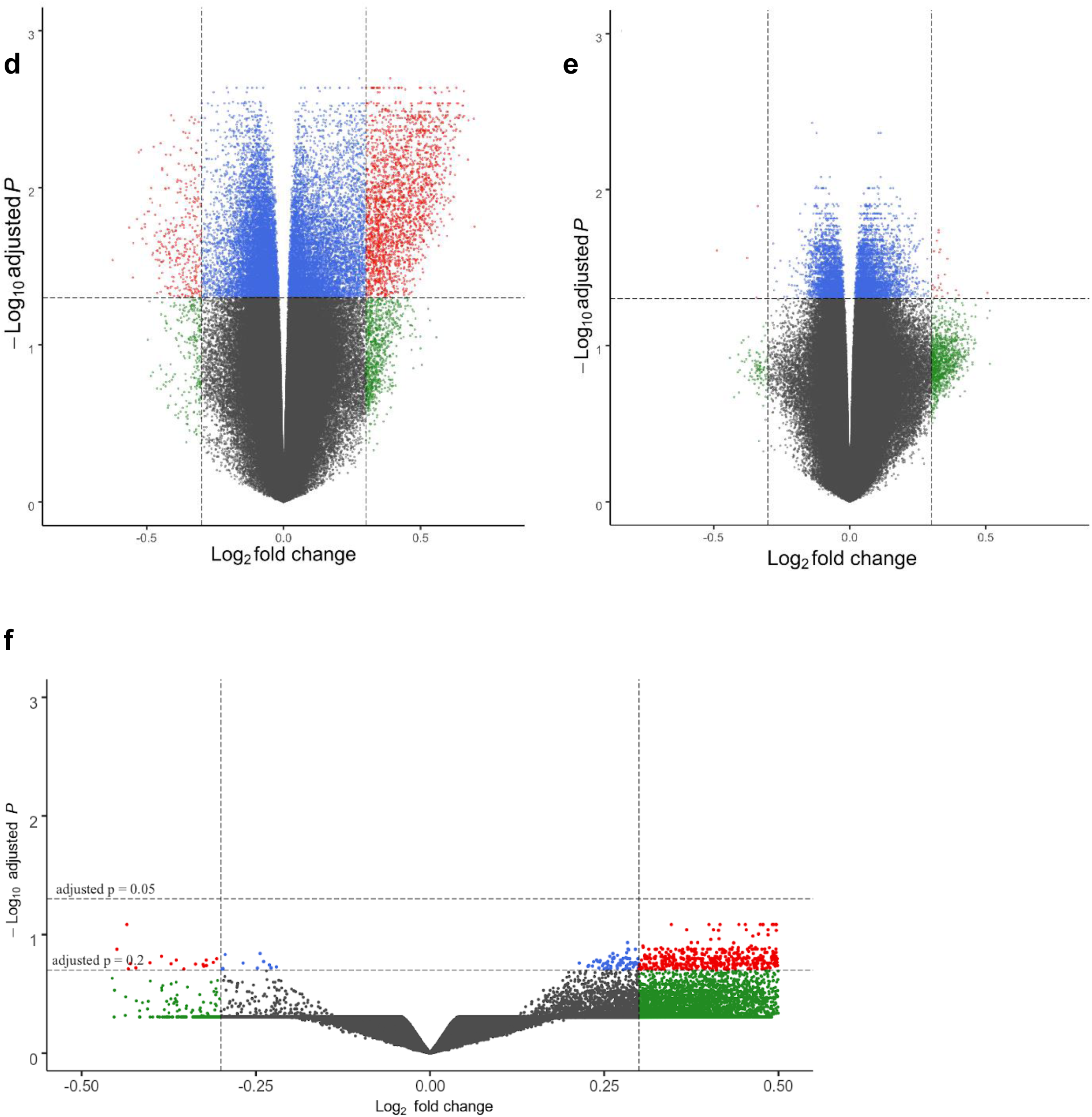
DNA methylome analyses of immune cells from TB-exposed individuals. Dendrograms of the unsupervised hierarchical clustering of the DNAm β-values of from **a**. HLA-DR, **b**. CD3 and **c**. PBMCs. “Con”: green=controls, “Exp”: purple=TB-exposed, red=TB index case. The scale defines the clustering Euclidean distance. Volcano plots of DMGs from **d**. HLA-DR, **e**. CD3 and **f**. PBMCs. Red dots represent DMGs above cut-offs (±0.3 Log_2_ fold change and BH-corrected *p*-value < 0.05, <0.1 or 0.2 as indicated).

Next, we identified the differentially methylated CpG sites (DMCs) and differentially methylated genes (DMGs) by comparing the TB-exposed and control groups for each cell population. To filter out the most significantly altered DMGs in the dataset, the stringency criteria of log_2_>|0.3| fold increased or decreased β-values and Benjamini-Hochberg (BH)-corrected *p*-value <0.05 (HLA-DR), <0.1 (CD3) and <0.2 (PBMC) were applied. The results are depicted as volcano plots, which show that DNA methylomes of TB-exposed most strongly differ in the HLA-DR cells as compared to control subjects, followed by the CD3 population, whereas PBMC datasets reveled fewer DMGs (Fig. 2d, e, f, Table 2). To highlight the locus position of the DMGs, chromosome maps were constructed (Suppl. Fig. S1a-c). Using the same stringency criteria as for the HLA-DR analysis, we tested whether DMGs would emerge when the datasets were arranged in other possible groups as derived from the demographics (>/< median age, sex, IGRA status). Neither age nor IGRA status generated any significant DMGs with these settings, and gender rendered three DMGs (X and Y chromosomes were removed in the initial filtering prior the analysis) in the HLA-DR positive and in the CD3 positive cells (Table 2).

**Table 2:** Differentially methylated genes (DMGs) identified through comparison of different study participant characteristics (TB-exposure, age, gender, IGRA status and BMI) in the three different cell populations, CD3+, HLA-DR+ and PBMC. DMGs were only identified when comparing the characteristic TB-exposure and gender.

| Characteristics |  | Cell type | DMGs | Gene name | p-value |
| --- | --- | --- | --- | --- | --- |
| **TB Exposure** | exposed vs. controls | HLA-DR | 9,786 | (not shown) | 0.05 |
|  |  | CD3 | 5,045 | (not shown) | 0.05 |
|  |  | PBMC | 496 | (not shown) | 0.2 |
| **Age** | < 30 vs. ≥ 30 | HLA-DR | 0 | | 0.05 |
|  |  | CD3 | 0 |  |  |
|  |  | PBMC | 0 |  |  |
| **Gender** | male vs.female | HLA-DR | 3 | GALNTL5;<br>MIR3201;<br>OPN3 |  |
|  |  | CD3 | 3 | COL9A1;<br>ACTG1;<br>REAT1E |  |
|  |  | PBMC | 0 |  |  |
| **IGRA** | positive vs.negative | HLA-DR | 0 |  |  |
|  |  | CD3 | 0 |  |  |
|  |  | PBMC | 0 |  |  |
| **BMI** | > 25 vs ≤ 25 | HLA-DR | 0 | | |
|  |  | CD3 | 0 |  |  |
|  |  | PBMC | 0 |  |  |

**Table 3:** Demographic data of the participants in the second recruitment (“test dataset”) ^†^ The standard deviation of the mean values of age, height, weight, and BMI.

| Characteristics | TB patients (n = 2) | TB-exposed (n = 2) |
| --- | --- | --- |
| Mean age (year)<sup>†</sup> | 19.5±1.5<sup>†</sup> | 35.5±17.5 |
| Mean height (cm)<sup>†</sup> | 176±7 | 171±3 |
| Mean weight (kg)<sup>†</sup> | 62.5±6.5 | 80±10 |
| Mean Body Mass Index (BMI)<sup>†</sup> | 19.9±0.6 | 27.3±2.5 |
| Sex (male/female) | 2/0 | 1/1 |
| Smoking (current/previous/never) | 2/0/0 | 1/0/1 |
| BCG (yes/no) | 0/2 | 2/0 |

### Functional enrichment analysis reveals common and unique interactomes in the datasets

Using the PANTHER Database, we investigated whether the identified DMGs were enriched in known pathways (Fig. 3a,b,c). The analysis revealed enrichment in pathways with relevance for TB infection, including hypoxia-inducible factor (HIF)1-α activation, Vitamin D metabolism and p38, Wnt, Notch, interleukin, chemokine, and cytokine signaling pathways^10–17^. Common pathways shared between at least two of the cell populations included B cell activation, glycolysis, angiotensin II signaling, and cholecystokinin signaling. Notably, several pathways named after their known functions in the nervous system were enriched in the studied cell populations, including pathways involved in axon guidance and adrenaline, acetylcholine and glutamate signaling. In the PBMC population but not in the lung cell populations, the interferon-γ signaling pathway was identified among the enriched pathways.

**Figure 3.**
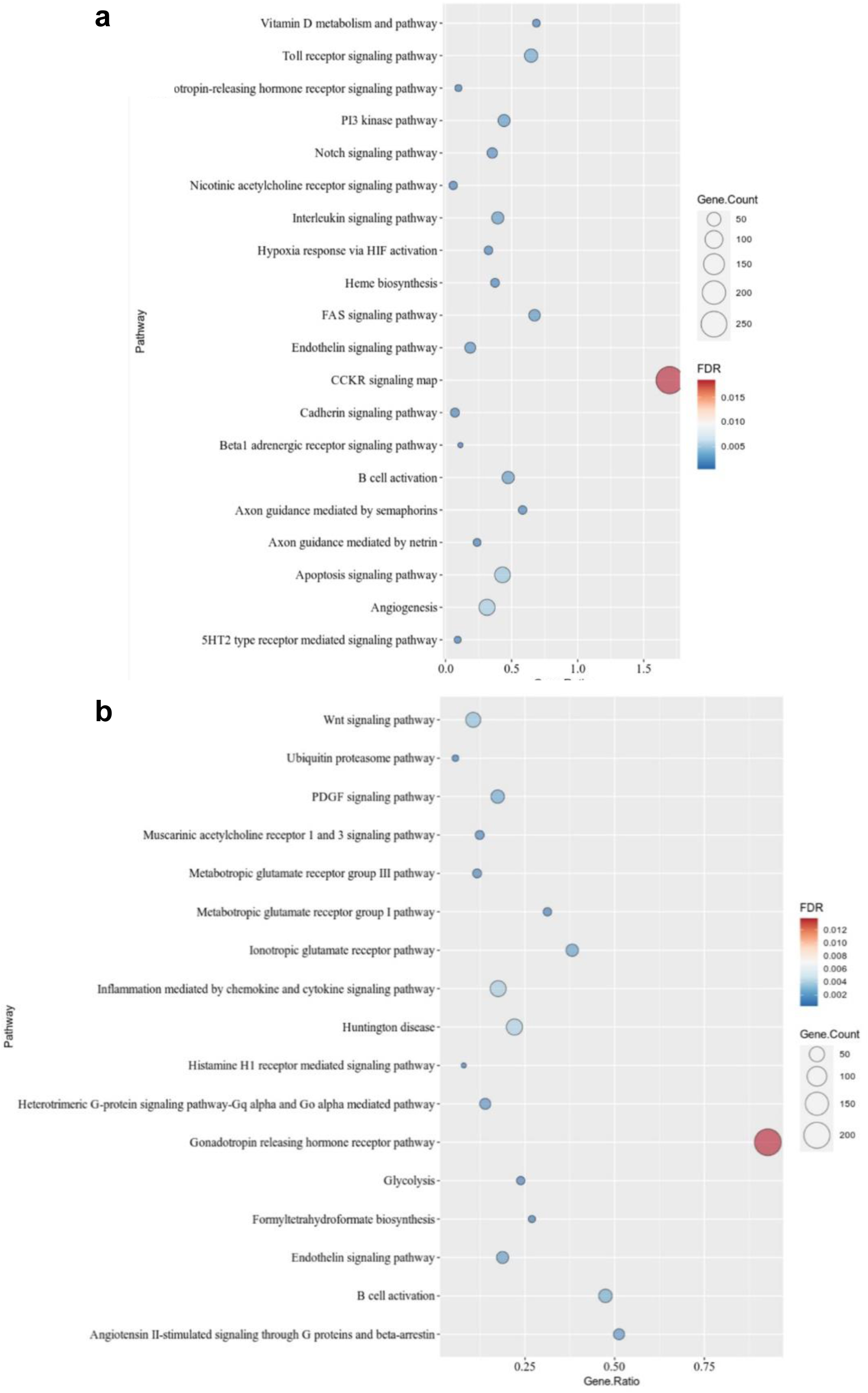
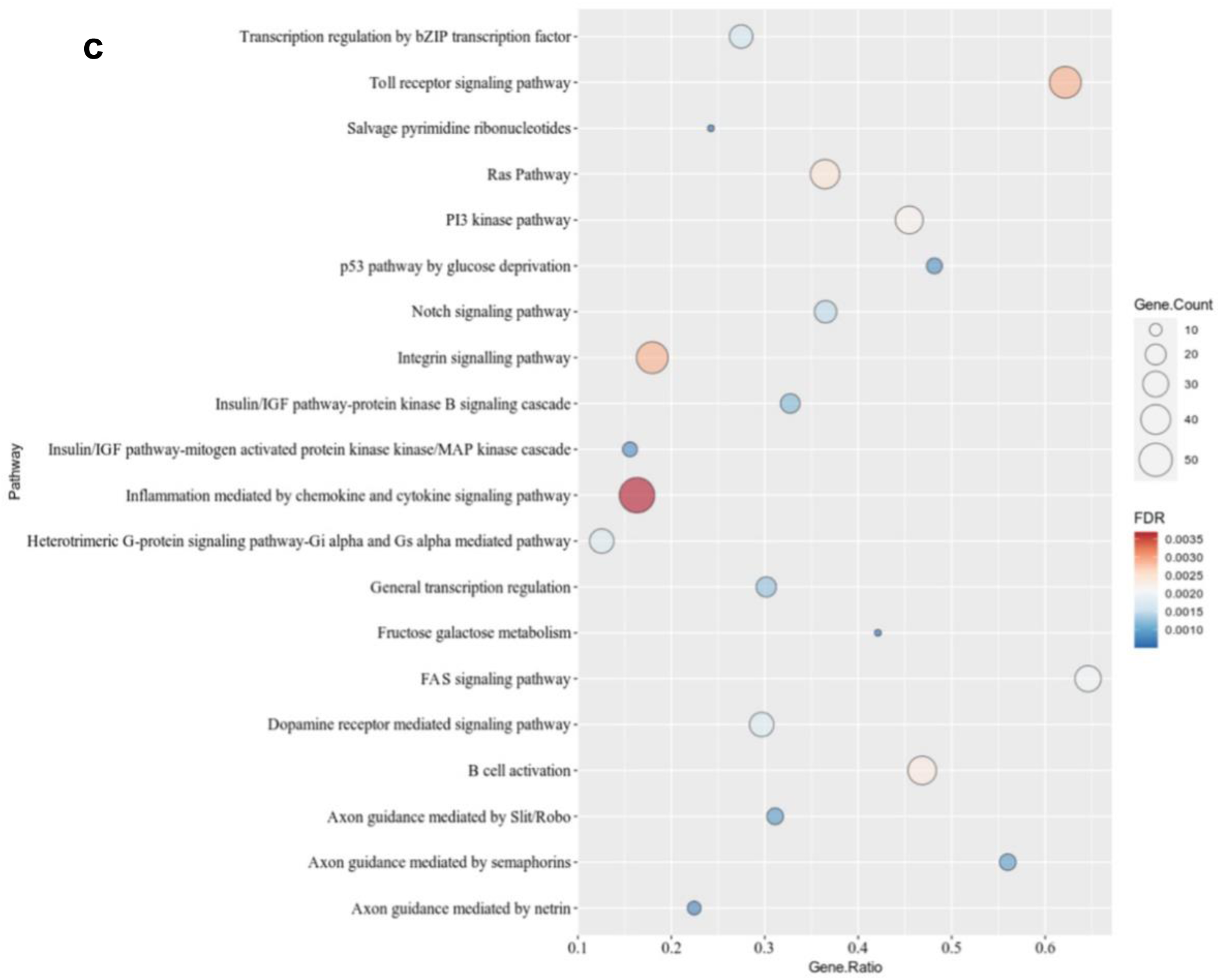
PANTHER pathway analysis of the identified DMGs with the cut-offs for the different cell populations given in Figure 2. Dot plots show the gene ratio, gene counts and FDR-corrected *p*-value for **a**. HLA-DR (top 20 pathways), **b**. CD3 (total 17 pathways), **c**. PBMC (top 20 pathways).

### Comparisons across cell populations and species reveals the existence of a common DNA methylome-based biosignature in mycobacteria-exposed immune cells

Given the fact that the interaction between mycobacteria and eukaryotes is evolutionary ancient, we predicted that highly conserved pathways exist that are common among the studied cell populations. Comparing the identified DMGs from the HLA-DR, CD3 and PBMCs in a Venn analysis, we discovered 185 common DMGs (Fig. 4a). We expanded the Venn analyses to include data from our previous work on BCG vaccine-induced DMGs that correlated with enhanced mycobacterial control^4^, as natural exposure to TB and BCG vaccination both represent *in vivo* encounters between mycobacteria and host immune cells. Even though the routes of mycobacterial exposure differ profoundly in these settings, a set of 151 DMGs could be identified as overlapping between our previous BCG study and all cell populations studied here (Fig. 4b), suggesting that a highly conserved epigenetic response to mycobacterial challenge exists.

**Figure 4:**
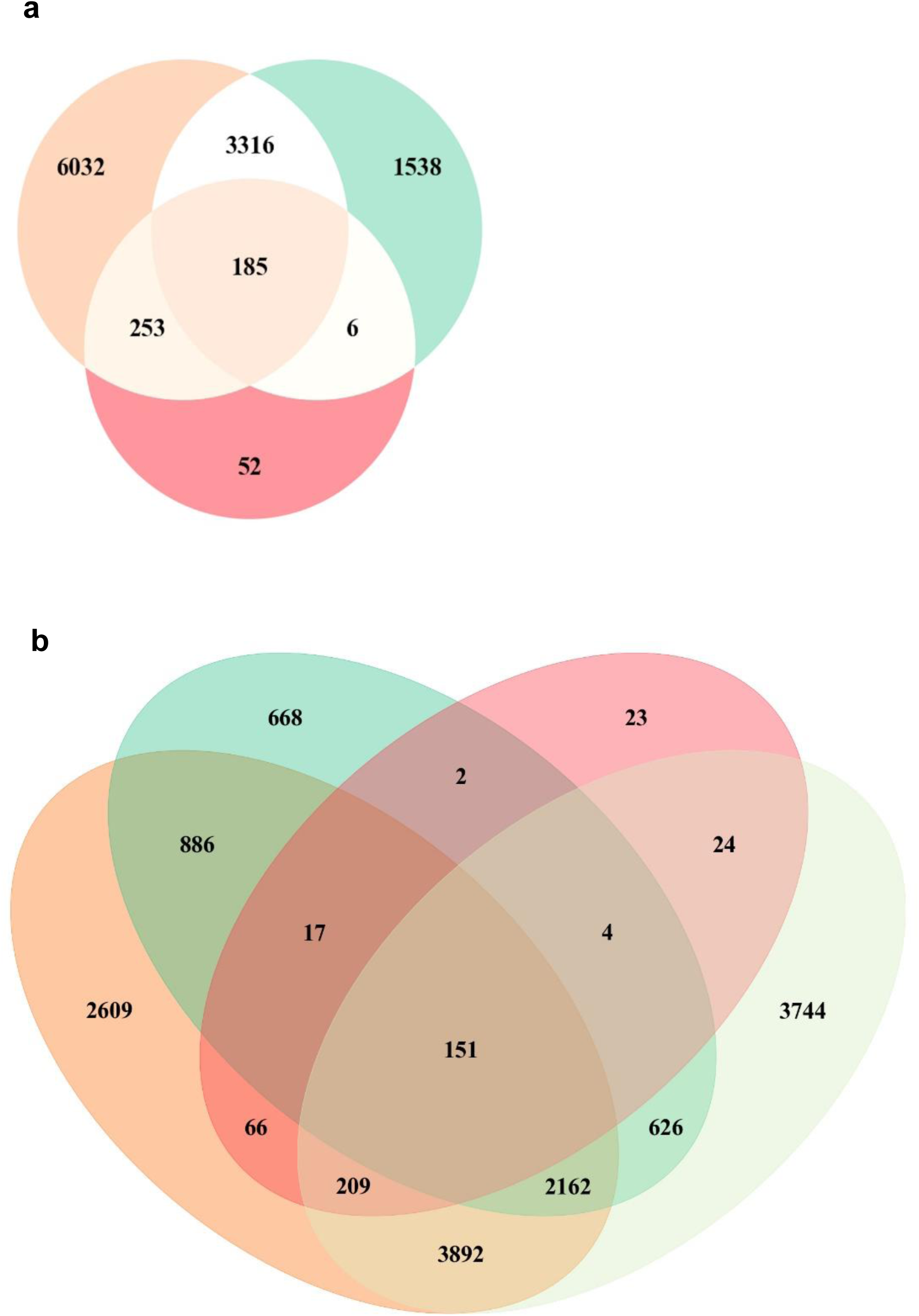
Venn analyses comparing DMGs, pathways and GO terms between different datasets. **a**. Overlapping DMGs derived from the HLA-DR (orange), CD3 (green) and PBMC (red) DNA methylomes. **b**. Overlapping DMGs from this study and from our previous work on BCG induced DMGs (light green).

In 2018, Hasso-Agopsowicz *et al*. described alterations in DNAm patterns in PBMCs from BCG-vaccinated individuals, with concomitant enrichment in many immune-related pathways^5^. In order to compare that study with ours, we performed PANTHER analysis with the 185 common DMGs and matched the identified enriched pathways, revealing that 75% of those pathways were the same as in the present study (Fig. 5a), further corroborating the relationship of the altered DNAm patterns induced through TB exposure and BCG-induced changes. In a recent study by Kaufmann *et al*., BCG-induced alterations of the epigenome in mice was correlated with protection against *M. tuberculosis* infection^6^. In order to translate our human DNA methylome signature to the signature identified in the mouse study, we searched for pathway overlaps between the two we performed a Gene Ontology (GO) enrichment analysis (Suppl. Fig. S2a-c). Figure 5b demonstrates that for our PBMC data, the GO terms “biological processes” overlapped 100% with the mouse study (same cell population) and to 31% and 65% for HLA-DR- and CD3-positive cells, respectively. In 2014, Saeed *et al* ^19^ demonstrated the induction of trained immunity pathways by another immune-training agent, β-glucan. We assessed possible pathway overlap with that study and although there were fewer overlaps as compared to the BCG-induced pathways described above. Again, the strongest correlation was found in the PBMC fraction, in this case in the GO terms “cellular components” (Fig. 5c and Table 1c).

**Figure 5.**
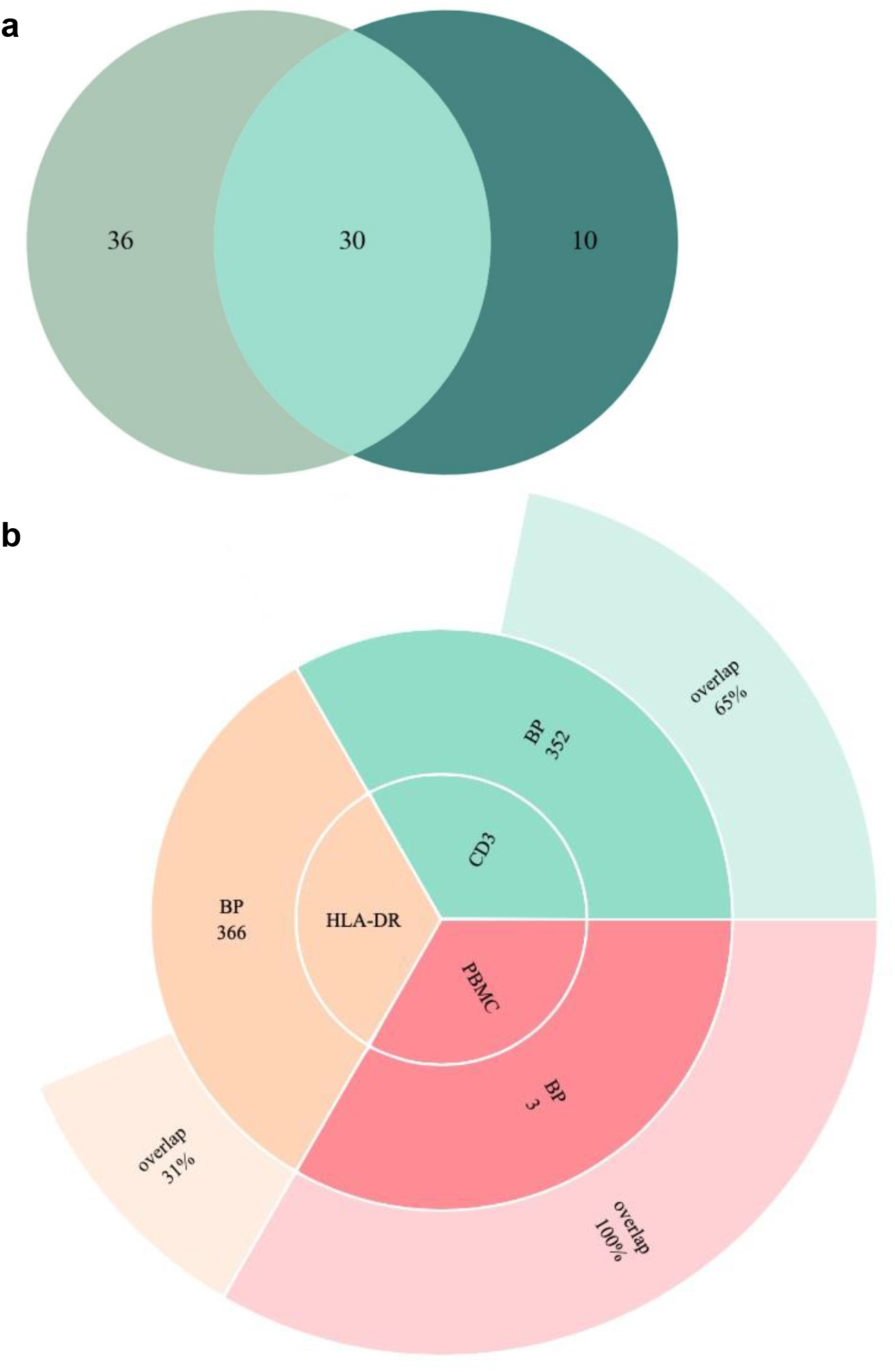
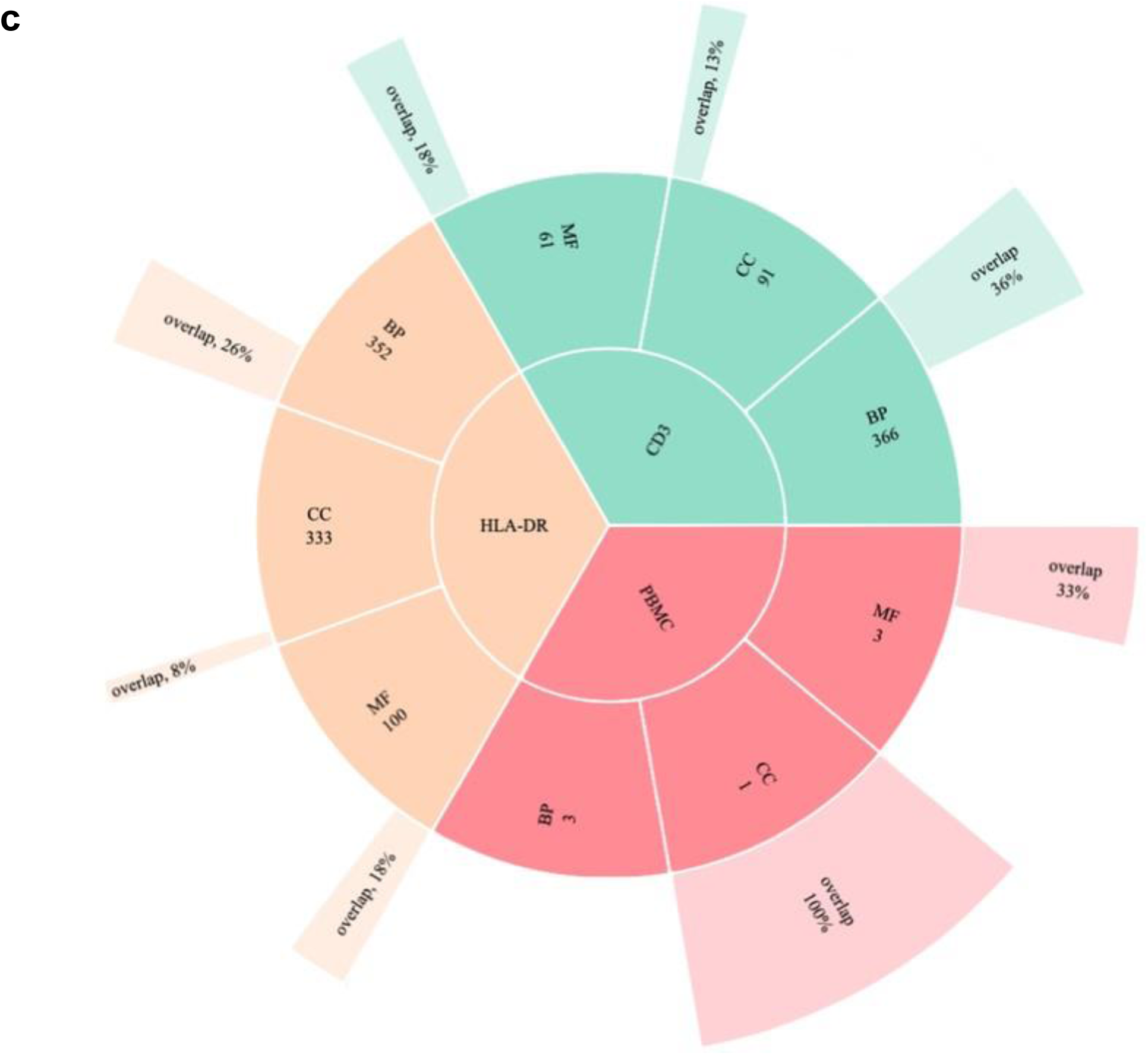
Pathway overlap with other studies’ results. **a**. Venn diagram describing the number of Panther pathways overlapping between the ones derived from the 185 common DMGs in this study (dark green) and Hasso-Agopsowicz *et al* (human BCG vaccine study, light green) **b**. Sunburst Plot describing the overlap of enriched GO biological processes emerging from a comparison between the GO data derived from the 185 common DMGs (Figure 4a) and Kaufmann et al (BCG study performed in mouse PBMCs). **c**. Sunburst Plot describing the overlap of enriched GO biological processes emerging from a comparison between the GO data derived from the 185 common DMGs and Saeed *et al* (study on trained immunity induced by β-glucan).

To validate how well the 284 CpG sites corresponding to the 185 overlapping DMGs performed in an unsupervised cluster analysis, we included one additional TB patient and two contacts, and collected HLA-DR-positive cells from induced sputum, as the DNA methylome data from this cell type was clearly outperforming the others with respect to accurate separation of the groups. Fig. 6 shows a *k* means-based dendrogram with a heatmap of the β values of the 284 CpG sites from both previous and new subjects’ DNAm data, revealing a distinct separation of the subjects in accordance with TB exposure.

**Figure 6.**
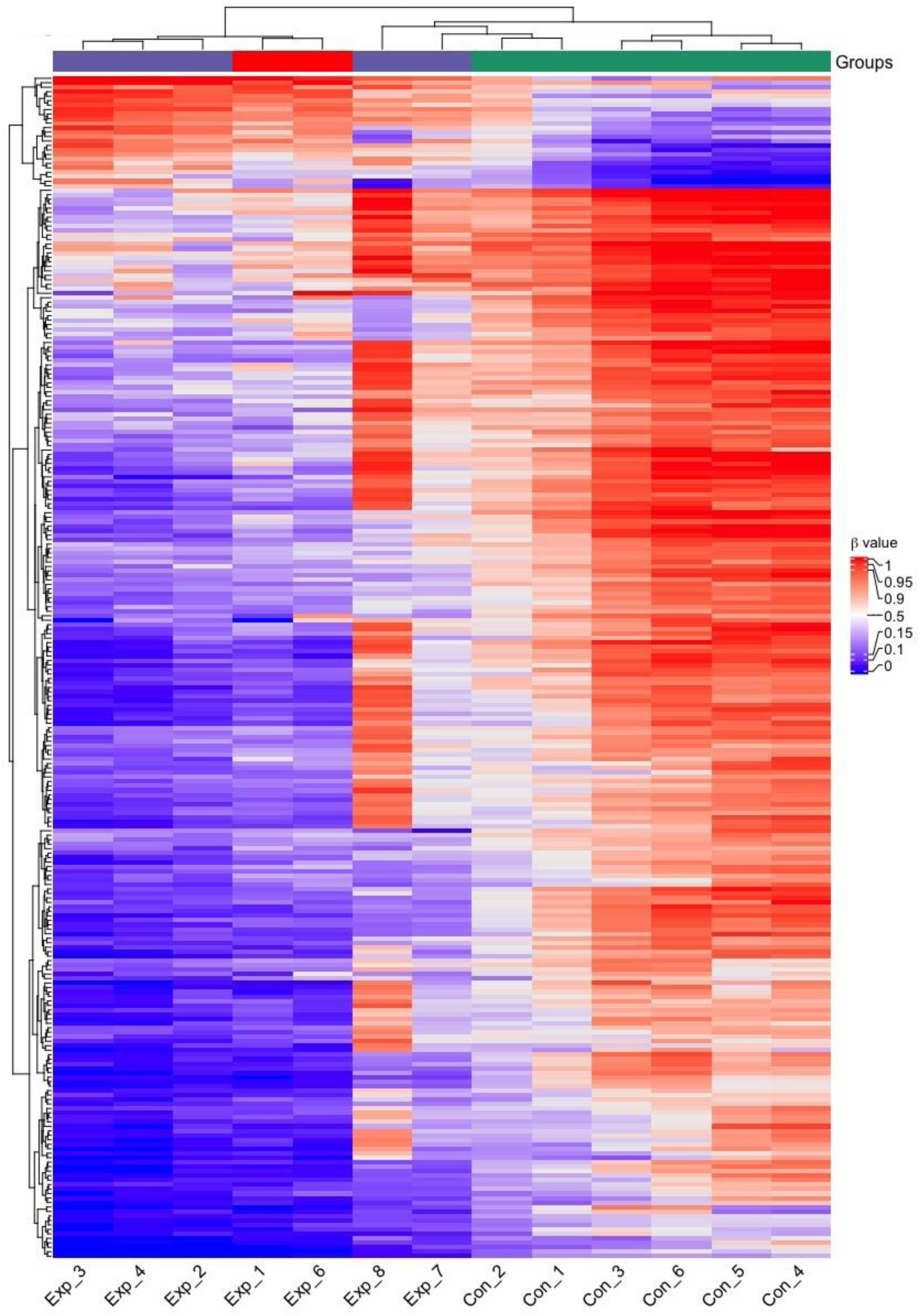
Heatmap of the HLA-DR-derived β values of the signature’s 284 CpG sites of the 6 initial subjects (Exp_1-4 and Con_1-6) and the three additional exposed subjects (Exp_6-8). Purple=exposed, red=TB index case, green=controls.

### In vitro BCG training of macrophages induced DNAm changes corresponding to exposure to TB

To investigate whether the BCG-induced epigenetic changes can be mimicked *in vitro*, we set up a BCG training experiment with macrophages isolated from donor blood that were trained with BCG and subsequently infected with the virulent *M. tuberculosis* strain H37Rv (expressing GFP) and monitored during the course of infection. In a subset of donors, BCG-trained cells displayed an increased capacity to kill *M. tuberculosis* (Fig. 7a-b) while cell viability was retained (Fig. 7c-d). DNAm analyses was performed on DNA isolated from these donors’ macrophages 24 hours after BCG training. We identified 7471 DMGs with the stringency criteria of DNAm difference > 30% and BH corrected *p*-value < 0.01. A PANTHER pathway analysis based on the identified DMGs showed significant enrichment in the Wnt signaling pathway, cadherin signaling pathways and angiogenesis, overlapping with the over-represented pathways in the HLA-DR+ and CD3+ cells from the TB-exposed individuals (Fig. 7e).

**Figure 7.**
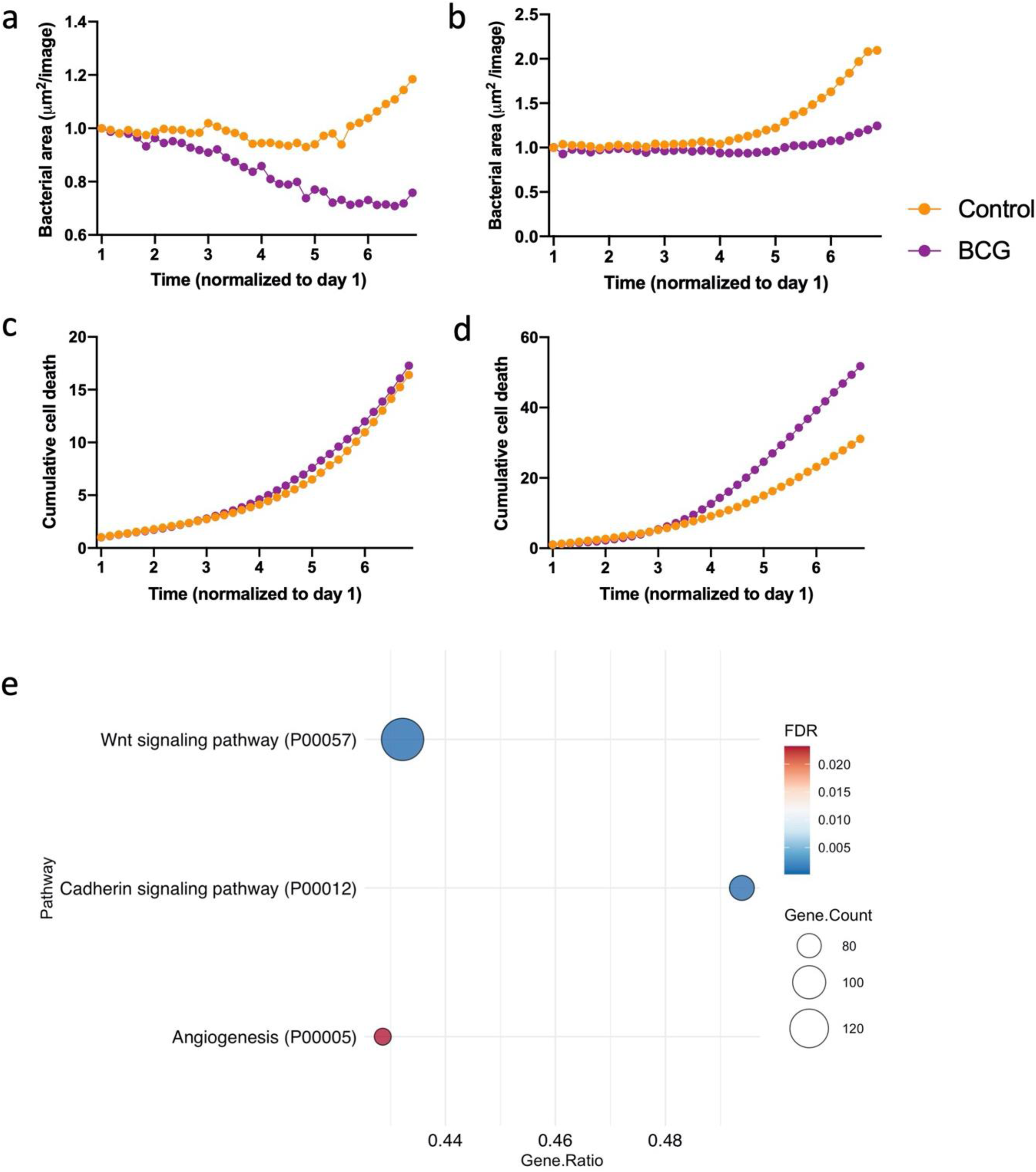
Macrophages were BCG-trained (purple) or left untreated (orange) before infection with *M. tuberculosis* (H37rv-GFP) or DNAm analysis. **a-b**. Total bacterial area (µm^2^/image) measured by green fluorescence over time. **c-d**. Cumulative cell death measured by the red fluorescent DRAQ7 DNA stain marking the nuclei of non-viable cells. **a-d** shows data from one donor separately with the median of two replicates per timepoint.**e**. PANTHER pathway analysis of the identified DMGs from DNA methylation analysis in the human primary macrophages exposed to BCG compared to controls. Dot plots show the gene ratio, gene counts and FDR-corrected *p*-value for 3 significantly over-represented pathways.

### BCG training of primary human macrophages induces morphological changes during M. tuberculosis infection

To better understand the mechanism by which BCG enhances the anti-mycobacterial activity of macrophages, we included N-glycolylated muramyldipeptide (MPD), alongside BCG in the training experiment with macrophages from ten donors. MDP is a component of the mycobacterial cell wall and has been described as the minimal component of BCG to induce trained immunity^20,21^. The infected macrophages were monitored in the IncuCyte^®^ S3 Live Cell Imaging system. We observed the emergence and expansion of a non-phagocytic cell type in BCG-trained macrophages from three donors (Fig. 8a-f and supplemental video S1), which seemed unaffected by high bacterial load. We termed this yet undefined cell type ‘corralling cells’, based on their apparent coordinated action to relocate and contain bacteria into focal points. We observed that distinct morphological changes occurred prior the emergence of the corralling cells and classified this process into 6 distinct stages shown in figure 8a-f. In staining experiments using Oil Red O, a lipid stain, in paraformaldehyde-fixed cells, we could demonstrate that the vacuole-like structures were not lipid droplets, excluding the possibility that this morphological state was reflective of foamy macrophages described previously ^22^ (Fig. 8g-h). The fixed cells were also stained with fluorescently conjugated antibodies (anti-α- smooth muscle actin (α-SMA), CD68, CD3, CD14, Collagen I and HLA-DR) to identify potential expression markers on the corralling cells. The HLA-DR antibody positively stained the macrophages, whereas the corralling cells were negative. (Fig 8i-j). No positive staining was observed for α-SMA, CD3, CD14 or collagen I (data not shown). Quantification of the number of corralling cells revealed that BCG-training cells but not MDP-training significantly induced the emergence of these cells (Fig. 9a). Finally, we evaluated the outcome for *M. tuberculosis* growth in wells with corralling cells. Figure 9b and supplemental video S2a (corralling) and S2b (non-corralling) show that in wells with corralling cells, the GFP-fluorescence of *M. tuberculosis* was strongly suppressed.

**Figure 8.**
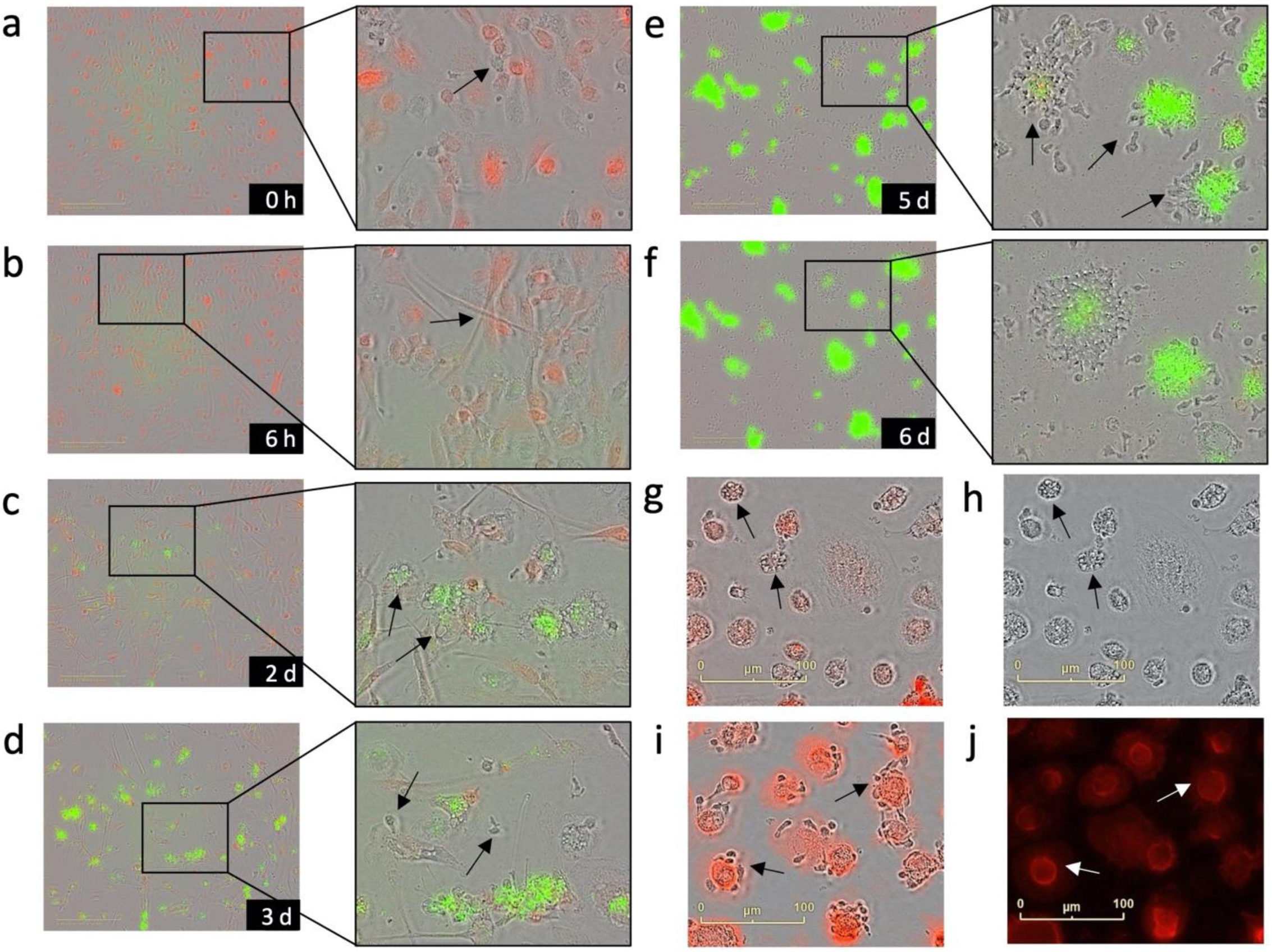
Morphological staging of the emergence of corralling cells (images representative of 3 donors with 4-6 technical replicates). **a**. Macrophages with red fluorescently labelled nuclei using Nuclight and *M. tuberculosis* (H37rv-GFP) fluorescent in green by GFP-expression, arrow showing intracellular *M. tuberculosis*. **b**. Stretching; Macrophages form extensions (100-300 µm long) during the first 24 hours **c**. Vacuolization; non-stretching macrophages become vacuolized (initially <1um in diameter, increasing to 15 um within 24-36 hours). **d**. Emergence; the presence of corralling cells is obvious at day 3 after which their numbers increase **e**. Corralling; corralling cells engage in pushing the bacteria into focal areas. **f**. Containment; *M. tuberculosis* bacteria are contained in the focal areas. **g-h**. Paraformaldehyde-fixed cells stained with Oil red O (red). Arrows indicate vacuolization. **i-j** Paraformaldehyde-fixed cells stained with fluorescently labelled anti-HLA-DR (red). Arrows indicate corralling cells.

**Figure 9.**
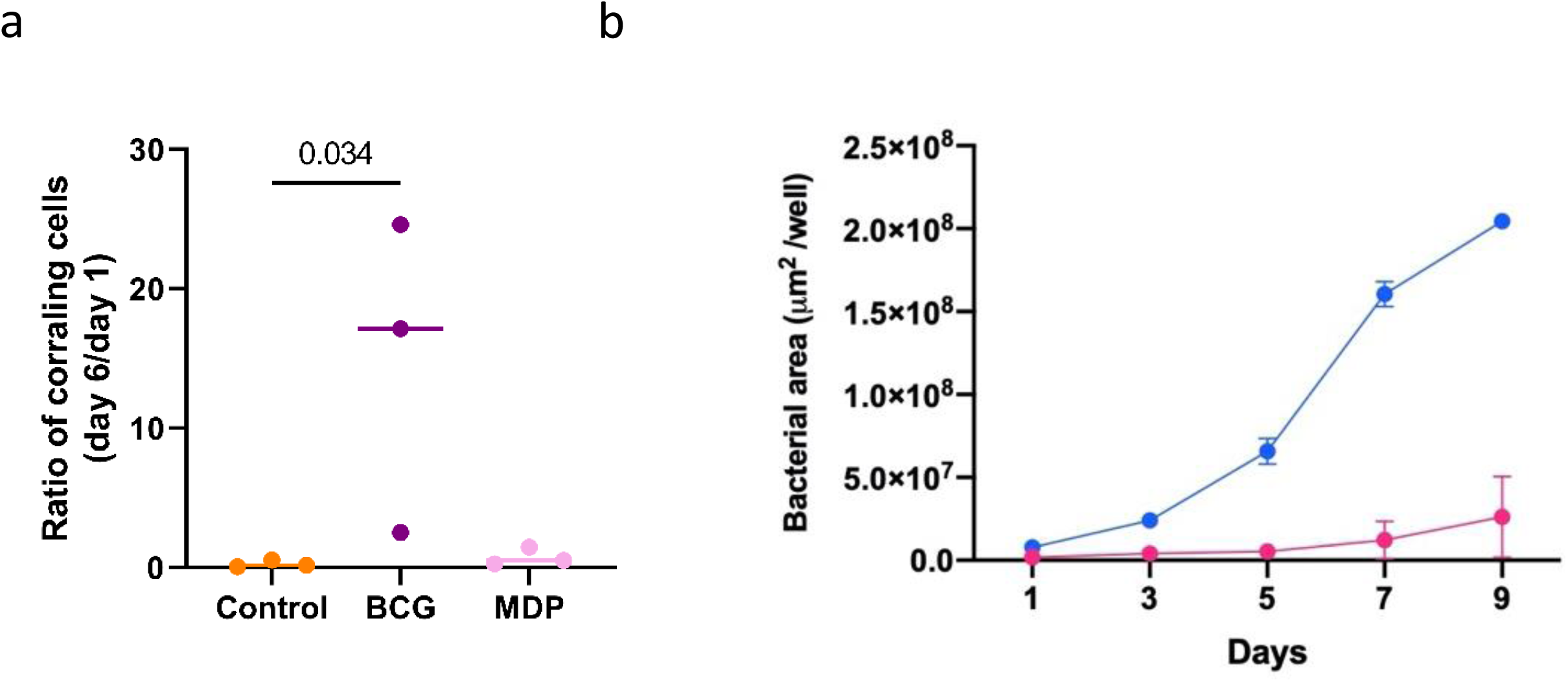
**a**. Quantification of corralling cells in *M. tuberculosis*-infected cultures with BCG- and MPD-trained macrophages. Cells with a size of 10-20 µm length were manually counted in 4-6 technical replicates from three donors. Statistical analysis was done with the Kruskal-Wallis test with Dunn’s multiple comparisons (*p*-value as indicated). **b**. *M. tuberculosis* (strain H37rv-GFP) fluorescence area (μm^2^/well) measured in 12-well plates in the IncuCyte_®_ S3 analysis tool for 2 donors with corralling cells (pink) and 2 donors without corralling cells (blue), error bars show SEM.

## Discussion

In this study, we present data suggesting that exposure to TB generates a distinct DNAm signature in pulmonary immune cells. The signature was found not only in those with active or latent TB infection, but also in individuals who are exposed but IGRA-negative. The finding that healthy, TB-exposed individuals also carry the signature opens the possibility that the epigenetic alterations reflect a host-beneficial reprogramming of the immune mechanisms rather than being induced by *M. tuberculosis* as a step to evade the immune defense. This notion is supported by our observation that the DMGs identified in the present study strongly overlapped with the epigenetic alterations identified in the *in vitro* BCG-trained macrophages, and the previously reported DNAm changes induced during BCG vaccination, which correlated with increased anti-mycobacterial capacity of macrophages^4^. In addition, we demonstrate that the GO data derived from our dataset display a strong overlap with data from a study on protective BCG vaccination in mice^6^.

BCG vaccination has convincingly been shown to induce heterologous immunity protecting against childhood mortality from other causes than TB^23,24^. Based on our finding that natural TB exposure and BCG vaccination trigger similar epigenetic changes we propose the hypothesis that a “beneficial exposure” to TB exists, which protects against other infections through heterologous immunity. Along the same line, it has been shown that a substantial fraction of individuals exposed to TB can be defined as ‘early clearers’, since they remain tuberculin skin test or IGRA negative^25^, suggesting effective eradication of the infection^25^. Identifying these early clearers and understanding the biology behind their resistance to TB infection could move the field forward towards novel strategies of TB prevention.

In concordance with the fact that macrophages constitute the main niche for mycobacterial replication, the strongest enrichment of DNAm changes was observed in the HLA-DR-positive cell population, which is dominated by alveolar macrophages. The pathways identified to be enriched in the HLA-DR-positive population have been described in the context of trained immunity, BCG exposure and TB. For example, activation of Hypoxia-Inducible Factor 1 α and glycolysis pathways (P00030 and P00024, respectively) are hallmarks of macrophages that have undergone the epigenetic changes reflective of trained immunity (reviewed in ^26,27^), which is induced in myeloid cells upon BCG-stimulation^21^. VEGF-release (P00056) by macrophages has been shown to recruit immune cells during granuloma formation^29^. Further, vitamin D has been shown to strengthen the anti-mycobacterial activity of macrophages^11,30^, and upregulation of components of the vitamin D pathway is linked to the production of anti-microbial peptides^12^, providing a possible effector mechanism for mycobacterial control. Recent literature on immune regulation through T cell-derived acetylcholine^31,32^ attributes relevance to the acetylcholine receptor pathway identified among the HLA-DR pathways.

Although macrophages and lymphocytes are not generally viewed as having many similarities, we found 34 of the identified pathways to overlap between HLA-DR- and CD3-positive cells. In data derived from the CD3 and PBMC populations, both of which represent lymphocytes, overlaps were identified for glycolysis, glutamate receptor and angiotensin II pathways. Interestingly, a metabolic shift towards increased glycolysis, representative of the Warburg effect, has been strongly associated with trained immunity^26^. However, the literature is dominated by the view that this event takes place in trained myeloid cells, while we identified this circuit in CD3 cells (lymphocytes) and not in the HLA-DR cells (dominated by macrophages). The glutamate receptor is widely expressed on immune cells and has been described as having an important regulatory role in T cells, which can also produce and release glutamate^33^. The role for angiotensin II pathway in TB remains elusive, while Angiotensin II Converting Enzyme 2 is currently in the spotlight due to fact that the SARS-CoV2 virus utilizes it as a receptor for entry into host cells^35^. In the PBMC population, which over all showed a weaker epigenetic response, we found the interferon-γsignaling pathway, which has a well-established role in anti-mycobacterial defense (reviewed in ^36^), among the reprogramed pathways.

Several studies have ascribed Wnt pathways immunomodulating functions and induction during *M. tuberculosis* infection (reviewed in ^14^) and M1 and M2 macrophages express distinct patterns of Wnt ligands. Here, in experiments with BCG-trained macrophage cultures, we observed the emergence of rapidly dividing cells with a size of approximately 1/4^th^ of the diameter of macrophages and negative for HLA-DR staining. A major limitation of the present study is that the corralling cells’ source of origin and mechanism for emergence could not be identified, but our ongoing studies are designed to characterize these cells. Taken together, we present data supportive of mycobacteria exposure induced DNAm changes that correlate with previous findings from studies on BCG vaccination including TB protection and trained immunity. We propose that the mycobacteria-specific DNAm changes promote the emergence of corralling cells to restrict mycobacterial growth, although this hypothesis needs further investigation.

## Methods

### Study design and participants

Patients with pulmonary TB, healthy participants with a history of TB-exposure and healthy controls, with an age ranging from 18 to 53 years, were enrolled at Linköping University Hospital and Linköping University, respectively. Included subjects (see Table 1 for demographics) donated peripheral blood and induced sputum samples^37^ (BioRxiv, pre-print, 2021, accepted for publication in *Epigenetics*) following oral and written informed consent (ethical approval obtained from the regional ethical review board in Linköping, #2016/237-31). The study protocol included questionnaires on respiratory and overall health, the evaluation of IGRA-status and sputum samples for DNA extraction. The subjects’ samples and questionnaires were not linked to any personal information at any stage of the study. For *in vitro* experiments, de-identified buffy coats were purchased from the blood facility at Linköping University Hospital. The buffy coats were obtained from healthy blood donors, who gave written consent of research use for the blood products.

### Induced sputum and pulmonary immune cell isolation

Induced sputum is a well-tolerated, non-invasive method to collect cells from the surface of the bronchial airways after inhalation of a hypertonic saline solution. The procedure of sputum induction takes approximately 30 minutes and is both cost effective and safe with minimal clinical risks^38^. Sputum specimens were collected as described by Alexis *et al* ^39^, with the following modifications: premedication with an adrenergic β2-agonist, salbutamol (Ventoline, 1ml 1mg/ml) was administrated before the inhalation of hypertonic saline, using a nebulizer (eFlow, PARI). The subsequent steps of sputum processing were adopted from Alexis et al. (2005)^40^ and Sikkeland *et al*^8^. The HLA-DR and CD3-positive cells were isolated using superparamagnetic beads coupled with anti-human CD3 and Pan Mouse IgG antibodies and HLA-DR/human MHC class II antibodies (Invitrogen Dynabeads, ThermoFisher, cat no 11041 and 14-9956-82, respectively). An initial positive selection was done with CD3 beads followed by a positive HLA-DR selection. Bead-coating and cell isolation were performed according to manufacturer’s protocol.

### PBMC isolation

PBMCs were isolated from whole blood (TB-exposed and controls) or from leukocyte rich fractions of blood obtained from healthy volunteers (Linköping University Hospital blood bank, Linköping). Isolation was performed by the method of density gradient centrifugation using Lymphoprep (Axis-Shield) and Sepmate-50 tubes (Stemcell Technologies) according to manufacturer’s protocol. IGRA status was determined on the whole-blood samples using QuantiFERON-TB Gold (Cellestis) following the manufacturer’s instructions.

### Cell culture and in vitro BCG training

Following PBMC isolation cells were seeded in cell culture treated flasks in Dulbecco’s Modified Eagle’s Medium (DMEM, Invitrogen) with 25 mM hepes, 100 U/ml penicillin and 100 μg/ml streptomycin (PEST, Gibco). Cells were incubated in 37 °C for 2 h before the non-adherent lymphocytes were washed away using warm Krebs-Ringer Glucose buffer (made in house). Complete DMEM supplemented with 10% pooled human serum, 2mM L-glutamine, 100 U/ml penicillin and 100 μg/ml streptomycin (all from Gibco) was then added to the cells along with immune training agents, or medium only as negative control for 24h. For the DNA methylation analysis and data presented in figure 7 a-d we used 10 μg/ml of BCG (freeze dried *M. bovis bacillus Calmette Guérin* Danish Strain). For the 10 donor experiment including MDP and data presented in figures 8 and 9 we instead used 3.33 μg/ml BCG or 3.33 μg/ml N-glycolylated muramyldipeptid (MDP) (Invivogen). Training agents were washed off and the cells were incubated for 6-7 days in complete medium with media change every 2-3 days. The cells were washed with PBS followed by trypsinization and reseeding of 5000 cells/ well in a 384-well plate (Corning Falcon) for infection experiments. DNAm analysis DNA was isolated at day 6 after training with BCG.

### M. tuberculosis culture and infection of macrophages

The laboratory *M. tuberculosis* strain, H37Rv, carrying the green fluorescent protein (GFP)-encoding pFPV2 plasmid (*M. tuberculosis*-GFP) was grown in Middlebrook 7H9 broth supplemented with albumin-dextrose-catalase (ADC, Becton-Dickinson), 0.05% Tween-80, and the selection antibiotic Kanamycin at 20µg/ml for two to three weeks at 37°C. The bacteria were then re-seeded in fresh broth for an additional 7 days to reach early log phase for use in experiments. For infection, the harvested bacteria were washed, resuspended in antibiotic-free complete DMEM and filtered through a Milliex-®SV 5.0µm syringe filter (Merck) to remove bacterial clumps. Macrophages were infected with *M. tuberculosis* at a multiplicity of infection (MOI) 0.5 and the cells imaged using an IncuCyte^®^ S3 (IncuCyte^®^ Live-Cell 120 Analysis System, Sartorius). The relative fluorescence signal (RFU) of bacteria was measured every 4 hours (20x, 4 images/well) until day 7 post-infection, analyzed using IncuCyte^®^ S3 software and expressed as total integrated intensity of green objects (GCUxμm^2^/image).

### Identification of morphological changes and quantification of corralling

Manual identification of morphological stages was performed by importing 8-bit grayscale images from the IncuCyte^®^ S3 software to ImageJ2^41^ using the Fiji package^42^. All images were processed to enhance contrast by 2% for ease of viewing. The number of coralling cells was manually estimated in images displayed in the IncuCyte^®^ S3 software. Bacterial area was determined using the basic analyzer tool in the IncuCyte^®^ S3 software.

### Immunohistochemistry for characterization of corralling cells

The 384-well culturing plates that were used to monitor the cells in the IncyCyte^®^ were fixed on day 10-11 from the infection. The plates were washed 2x in PBS before adding 4% paraformaldehyde in PBS for 30 minutes. The plate was washed again x2 in PBS and stored in PBS in 4 degrees until use. For the immunohistochemistry, the wells stained with intracellular markers (α-SMA) were permeabilized with 50 ul PBS (made in house) containing 1% bovine serum albumin (BSA) (Sigma-Aldrich, St. Luis, Missouri, U.S) and 0.2% Triton X-100 (Sigma Aldrich)/well for 10 min. No permeabilization was used for the staining of extracellular markers (CD3, CD14, CD68, HLA-DR, Collagen α2 Type I). The wells were washed 3 times with PBS and then 50 ul blocking buffer containing 1% BSA in PBS was added and the plate was incubated at RT for 1 h. The antibodies were diluted in PBS 1% BSA and added in a volume of 10 ul/well. The antibodies used were CD68 Alexa Fluor 647 diluted 1:25 (Cat no: sc-17832, Santa Cruz Biotechnology, Dallas, Texas, U.S), CD68 Alexa Fluor 647 diluted 1:20 (Cat no: 562111, BD biosciences, Franklin Lakes, New Jersey, U.S), α-SMA Alexa Fluor 488 diluted 1:50 (Cat no: sc-32251, Santa Cruz Biotechnology), CD3 FITC diluted 1:50 (Cat no: 349201, BD Biosciences), CD3 FITC diluted 1:50 (Cat no: 300306, Biolegend, San Diego, California, U.S), HLA-DR PE diluted 1:20 (Cat no: 327007, Biolegend), CD14 FITC diluted 1:20 (Cat no: 345784, BD biosciences), Collagen α2 Type I Alexa Fluor 647 diluted 1:50 (Cat no: 393573, Santa Cruz Biotechnology). For isotype controls we used normal mouse isotype control IgG_1_ Alexa Fluor 647 (Cat no: sc-24636, Santa Cruz Biotechnology) diluted 1:50, normal mouse isotype control IgG_2a_ Alexa Fluor 488 (Cat no: sc-3891, Santa Cruz Biotechnology) diluted 1:50) and normal mouse isotype control IgG_2b_ FITC (Cat no: 0090480, BD Biosciences). The plate was incubated for 1 h in RT. The pate was 3x in PBS and 20 ul Dako Mounting Medium (Agilent, Santa Clara, California, U.S) was added. The plate was read in the IncuCyte^®^ S3 Live Cell Imaging system at x20.

### DNAm sequencing and data analysis

DNA from PBMCs, HLA-DR-positive, CD3-positive cells, and the cultured human primary macrophages was extracted using the AllPrep DNA/RNA Mini Kit (Qiagen, Hilden, Germany) according to the manufacturer’s instructions. Genome-wide DNAm analysis was performed using the HumanMethylation450K BeadChip (Illumina, USA) array (for the HLA-DR, CD3, PBMC samples) and reduced representation bisulfite sequencing (Diagenode’s RRBS) read in Illumina’s HiSeq 2000 (for the cultured human primary macrophages) at the Bioinformatics and Expression Analysis (BEA) Core Facility at Karolinska Institute, Stockholm. The methylation profiles from the HumanMethylation450K BeadChip analysis for each cell type were analyzed from the raw IDAT files in R (v4.0.2) using the *minfi* (v1.36.0) with subset-quantile within array (SWAN) normalization^43,44^ and *ChAMP* (v2.19.3) with beta-mixture quantile normalization (BMIQ) packages^45,46^. The type I and type II probes were normalized using the quantile normalization method. Using the default setup of the *ChAMP* package, following probes were filtered out: i) probes below the detection *p*-value (>0.01), ii) non-CpG probes, iii) multi-hit probes, and iv) all probes of X and Y chromosomes. Cell type heterogeneity was corrected for the PBMC cell types using the Houseman algorithm^47^ and batch effects were fixed using *ComBat* from the *SVA* package (v3.38.0). Differential methylation analysis was performed with the linear modeling (lmFit) using the *limma* package^48^ (v3.46.0) in a contrast matrix of the TB-exposed and TB-non-exposed (Control) individuals. All Differentially methylated CpGs (DMCs) were considered significant at the Bonferroni-Hochberg (BH) corrected *p*-value < 0.05 (for HLA-DR cell types), <0.1 (for CD3 cell types) and <0.2 (for PBMC cell types). The DNA from the human primary macrophages was sequenced using the Diagenode’s RRBS due to a lower DNA yield. The sequenced reads were quality checked using the FastQC^49^ (v0.11.9). The sequences were trimmed to remove artificially filled-in cytosines at the 3′ end using the TrimGalore (v.0.6.5)(https://github.com/FelixKrueger/TrimGalore) with a phred score cutoff of 20 and quality checked again after trimming. The trimmed sequences were aligned with the human reference genome (hg38.13) using Bowtie2^50^ and removed the duplicates using the Bismark (v.0.22.3)^51^.

The methylation extractor from Bismark was used to extract the CpG methylation data from the sequences. The SAMtools (v1.7) package^52^ was used to sort the binary alignment files (BAM) files on CpG-site chromosomal location and converted to sequence alignment map (SAM) files. The methylated and unmethylated CpG counts were extracted and combined using the DMRfinder^53^ (v0.3) package in R (v4.0.2). The CpG-sites located in the X and Y chromosome as well as CpG-sites from mitochondrial DNA were filtered out. For differential methylation analysis the *methylKit*^54^ (v1.18.0) package was used. CpG-sites with a read coverage > 10 with both methylated and unmethylated reads were removed from the analysis. The function “calculateDiffMeth” from methylKit package using logistic regression was used as statistical test with “before” and “after” BCG-training as treatment used to identify differentially methylated CpG-sites DMCs. The DMCs were annotated to the official gene symbol using *org*.*Hs*.*eg*.*db* (v3.12) and AnnotationDbi (v1.52) packages using the human genome version hg38. DMGs were defined as sited with a methylation difference of 30% between the conditions and an BH corrected *p*-value of <0.01.

### Unsupervised cluster analysis

Hierarchical clustering of the all TB-exposed and control individuals was performed with the normalized β-values obtained after the data filtration in each cell type individually. The distance was calculated using the Euclidean distance matrix. The *dendextend*^55^ (v1.14.0) and *ape*^56^ (v5.4-1) packages in R were used to construct the horizontal hierarchical plots from the three different cell populations using the *hclust* and *dendrogram* functions.

### Structural annotations

The *EnhancedVolcano* package^57^ (v1.8.0) was used to generate the individual volcano plots from all cell populations. The *ChromoMap* package^58^ (v0.3) was used to annotate and visualize the genome-wide chromosomal distribution of the DMGs. The interactive plots were generated using the *plotly* (v4.9.3) package.

The heatmaps were generated from the filtered DMGs with their respective CpGs for each cell type using the *ComplexHeatmap* (v2.6.2) package^59^. The clustering dendrogram in heatmaps were plotted using the Euclidean distance matrix.

### Pathway and functional enrichment analyses

We used the PANTHER database (PantherDB v15, 16)^60^ to identify the enriched pathways related to our identified DMGs. In addition, to assess functional enrichment, we used the *ReactomePA* (v1.34.0) package^61^ with 1000 permutations and the BH-corrected *p*-values. Within the package, GO and Kyoto Encyclopedia of Genes and Genomes (KEGG) were used and using *clusterProfiler*^62^ (v3.18.1), we performed KEGG pathway^63^ enrichment analysis (data not shown). To enhance the visualization and better understanding of the enrichment result, *GOplot*^64^ (v1.0.2), another package was used. The pathway enrichment was also calculated using the topology-based ontology methods using *RontoTools*^65^ (v2.18.0), *SPIA*^67^ (v2.42.0) and *pathview*^68^ (v1.30.1) was used to visualize the related pathways with the KEGG pathway maps (data not shown).

### Venn and overlap analyses

Venn analyses were performed to detect the DMGs overlapping between cell populations and between studies. We constructed the Venn diagrams by using *matplotlib-venn* package (https://github.com/konstantint/matplotlib-venn) using in-house python script. The overlap analyses were calculated and plotted using the *go*.*Sunburst* function from plotly using an in-house python script.

### Statistical analyses

All differences with a *p-*value < 0.05 were considered significant if not otherwise stated. We calculated family-wise error rate (FWER) using the BH correction method. All analyses were performed in R (v4.0.2) with the mentioned packages.

## Data Availability

The data will be available for research after final publication of the manuscript upon reasonable request.

## Acknowledgements

This study was funded through generous grants from Forskningsrådet Sydöstra Sverige (FORSS-932096), the Swedish Research Council (2015-02593 and 2018-02961) and the Swedish Heart Lung Foundation (20150709 and 20180613). J.D is a postdoctoral fellow supported through the Medical Infection and Inflammation Center (MIIC) at Linköping University. We direct our gratitude to the staff at Linköping University Hospital and the Vrinnevi Hospital in Norrköping for assistance in sample collection and all the subjects for donating samples. The DNA methylome data were generated at the Bioinformatics and Expression Analysis Core Facility at the Department of Biosciences and Nutrition, which is supported by the Board of Research at the Karolinska Institute, Stockholm. The computations were enabled by resources provided by the Swedish National Infrastructure for Computing (SNIC) at Linköping University campus partially funded by the Swedish Research Council through grant agreement no. 2018-05973.

## Author contributions

M.L., N.I., J.P. and C.B. designed the study, I.P., N.I., C.B., L.K., B.A. and E.K.D performed the laboratory work and the related analyses, J.D. and M.L. designed and performed the bioinformatic analyses of the data, J.D. and L.K wrote the scripts for analysis and created figures. N.I., I.P., J.D., L.K., C.B., E.K.D. and M.L wrote the manuscript. J.D and M.L are co-authors of a patent application “Biomarker for detection of mycobacterial infection and exposure” filed on February 2^nd^ with the Swedish Patent Registry (#100692).

