## Supplementary figures and images for "DNA methylation profiling of immune cells from tuberculosis-exposed individuals overlaps with BCG-induced epigenetic changes and correlates with the emergence of anti-mycobacterial ‘corralling cells’"

### Suppl. figure 1a

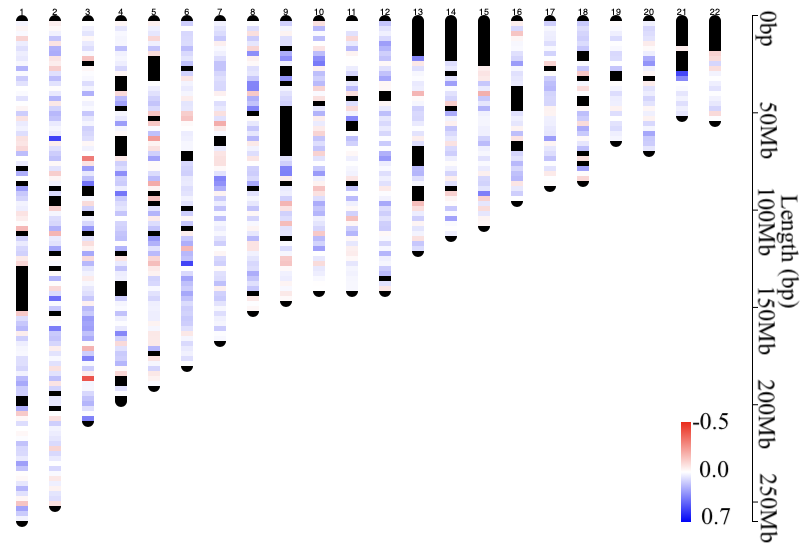

### Suppl. figure 1b

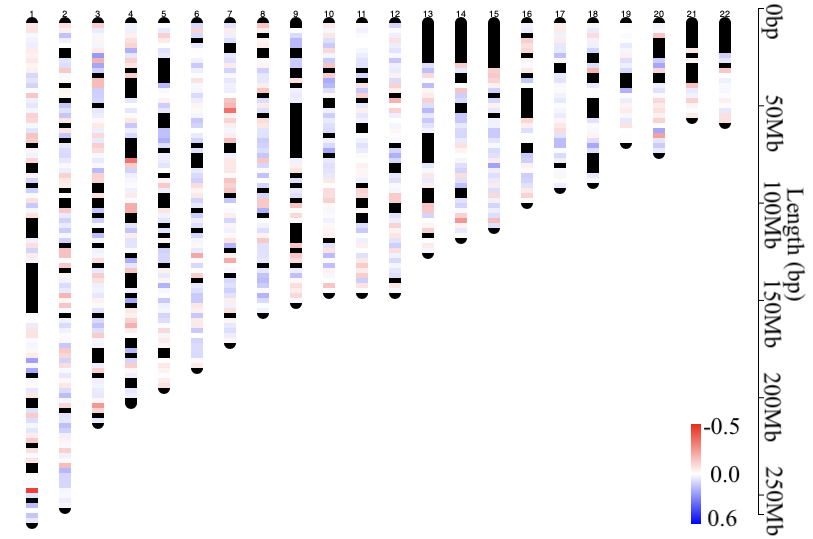

### Suppl. figure 1c

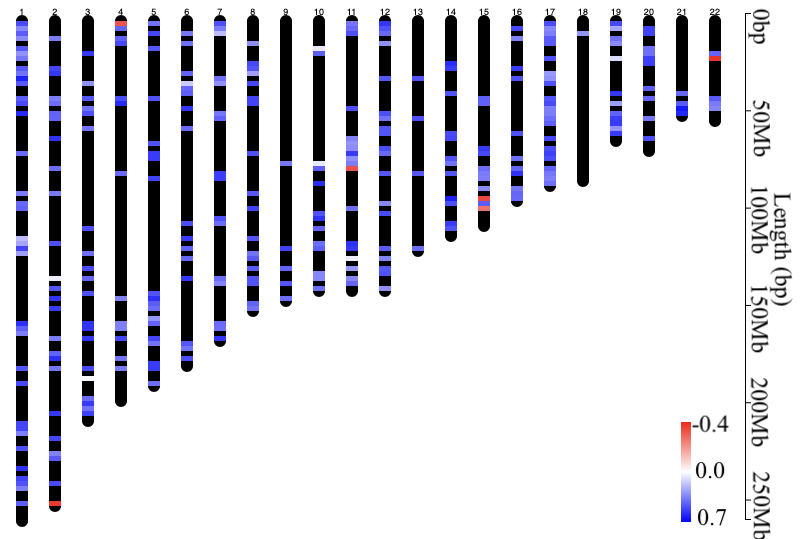
